# The impact of cognitive ability on multitalker speech perception in neurodivergent individuals

**DOI:** 10.1101/2022.09.19.22280007

**Authors:** Bonnie K. Lau, Katherine Emmons, Ross K. Maddox, Annette Estes, Stephen Dager, Susan J. (Astley) Hemingway, Adrian KC Lee

## Abstract

The ability to selectively attend to one talker in the presence of competing talkers is crucial to communication. Here we investigate whether cognitive deficits in the absences of hearing loss can impair speech perception. We tested typical hearing, neurodivergent adolescents/adults with autism spectrum disorder, fetal alcohol spectrum disorder, and an age- and sex-matched neurotypical group. We found a strong correlation between IQ and speech perception, with individuals with lower IQ scores having worse speech thresholds. These results demonstrate that deficits in cognitive ability, despite intact peripheral encoding, can impair listening under complex conditions. These findings have important implications for conceptual models of speech perception and for audiological services to improve communication in real-world environments for neurodivergent individuals.

---

Multitalker speech perception, the ability to selectively attend to one talker in the presence of several competing talkers, is a crucial yet often underappreciated skill employed in everyday life. To purchase a drink in a loud coffee shop, one must selectively tune-in to the barista’s voice while filtering out the voices of other customers. To learn in a classroom, a student needs to ignore the voices of other students to focus on the teacher’s voice. In real-world settings involving many people talking at once, neurotypical listeners with typical hearing can effortlessly attend to the desired talker even when competing voices are significantly louder (Brungart, 2001). The ability to organize and extract meaning from competing sounds in these complex acoustic environments is often referred to as auditory scene analysis or the cocktail party problem (Bregman, 1990).

Past research has demonstrated that when hearing loss degrades the peripheral encoding of sounds, deficits in multitalker speech perception are observed (reviewed in Moore, 2003; Shinn-Cunningham & Best, 2008). There is evidence that peripheral deficits in sound encoding cause cascading deficits in later stages of processing, including segregating, grouping, and selectively attending to the incoming sound mixture (Dai et al., 2018; Grimault et al., 2001), as well as the processing of speech-specific information (Obler et al., 1991; Peelle et al., 2011). What is less clear is whether impairments in cognitive processes in the absence of hearing loss can also result in speech perception difficulty. Prior investigations assessing the aging auditory system provide some evidence that deficits in speech perception may be related to cognitive decline (Gallun & Best, 2020; Humes et al., 2013; Taljaard et al., 2016; Van Gerven & Guerreiro, 2016). However, a specific relationship between age-related changes in cognitive function and multitalker speech perception deficits is difficult to establish because many older individuals experiencing cognitive decline also have hearing loss.

Successful auditory scene analysis relies first on the accurate encoding of the incoming sound mixture by the ear and auditory nerve fibers, followed by the integration of sounds between the ears (see Bronkhorst, 2015 for conceptual model). These spectro-temporal patterns encoded by the auditory periphery are then segregated and grouped into separate auditory streams based on acoustic features, such as pitch or spatial cues, that indicate where the sound is coming from (Best et al., 2006; Bizley & Cohen, 2013; Bronkhorst, 2000; Shinn-Cunningham et al., 2017). From there, more complex processing of the lexical, syntactic, and semantic information in the speech signal occurs (Davis & Johnsrude, 2007). Importantly, cognitive processes, including long and short-term memory, as well as attentional control, are thought to be required throughout both auditory grouping and complex processing stages (Michalka et al., 2015). The interplay between these cognitive and auditory perceptual processes are often modelled as complex functions, with involvement of both feedback and feedforward mechanisms (Bronkhorst, 2015). Moreover, there is evidence that the selective enhancement of relevant speech is influenced by an attention-mediated feedback loop (Ahveninen et al., 2011; Bronkhorst, 2015; Ding & Simon, 2012; O’Sullivan et al., 2015; Woldorff & Hillyard, 1991). Nonetheless, many aspects of the relationship between cognitive and auditory processes and how they function during multitalker speech perception are not fully understood.

This study investigated the effect of cognitive deficits on multitalker speech perception, in the absence of hearing loss. We tested neurodivergent adolescents/young adults with varying intellectual quotient (IQ) but clinically normal hearing thresholds who belonged to one of three diagnostic groups: autism spectrum disorder (ASD), fetal alcohol spectrum disorder (FASD), and an age- and sex-matched neurotypical comparison group (Comparison). We hypothesized that difficulty perceiving speech would be correlated with deficits in cognition in each of our participant groups.

ASD is a developmental condition characterized by difficulties with social communication and interactions, as well as restricted interests and repetitive behaviors (American Psychiatric Association, 2013). FASD describes a range of physical, functional, and neurological outcomes associated with prenatal alcohol exposure. These outcomes include physical differences such as facial dysmorphias as well as cognitive challenges, such as deficits in working memory, learning, communication, and attention (Astley, 2004, 2013).

There are many studies documenting self/caregiver reports of auditory processing differences, including hyper- and hypo-sensitivity to sound, aversions to sound, as well as difficulty perceiving speech under noisy conditions for individuals with ASD as well as FASD (Haesen et al., 2011; McLaughlin et al., 2019; O’Connor, 2012; Tomchek & Dunn, 2007). Difficulty understanding speech under noisy conditions is commonly reported by individuals with ASD in particular (e.g., Birch, 2003; Grandin, 2006). However, there are no prior studies measuring speech perception abilities in individuals with FASD and the few studies conducted in individuals with ASD report variable findings of whether speech perception deficits are indeed observed and if so, under what conditions (Alcántara et al., 2004; DePape et al., 2012; Groen et al., 2009; Schelinski & von Kriegstein, 2020). Variability in the results of these studies may be resulting from heterogeneity in the participants themselves; thus, motivating our current study to further consider the role of cognition in speech perception under complex conditions. Individuals with ASD and FASD show a range of cognitive ability, from above average IQ to intellectual disability, but how cognitive ability impacts speech perception is not well understood.

We took several characteristics of individuals with ASD and FASD into consideration in our study design and analyses. First, we conducted an audiometric screen at octave frequencies between 250-8000 Hz (<=20 dB Hearing Level), as well as an otoacoustic emissions screen, to ensure clinically normal hearing thresholds in all participants, as there is an increased incidence of hearing loss in both populations (Klin, 1993; McLaughlin et al., 2019; Rosenhall et al., 1999; Szymanski et al., 2012). Second, we chose a receptive speech perception task in which participants did not require verbal responses, as our intent was to assess speech perception rather than speech production. Finally, we chose an adaptive threshold task, so that task difficulty would be varied systematically based on individual performance. Importantly, an adaptive threshold task design ensured that at the start of each run, participants were able to perform the speech perception task when conditions were favorable.

## Methods

### Participants

Forty-nine subjects (*n* = 12 ASD group; *n* = 10 FASD group, *n* = 27 Comparison group) participated in the study (see Table 1 for demographic information). Twelve adults diagnosed with autism spectrum disorder (ASD) were recruited from a larger longitudinal study conducted at the University of Washington. The original cohort consisted of 72 children diagnosed with ASD between the ages of 3 and 4 years. Diagnoses of ASD, according to criteria from the fourth edition of the *Diagnostic and Statistical Manual of Mental Disorders* (American Psychiatric Association, 1994) were made by a licensed clinical psychologist or supervised graduate student using: 1) the Autism Diagnostic Interview-Revised (Lord et al., 1994), 2) the Autism Diagnostic Observation Schedule (ADOS; Lord et al., 1989), 3) medical and family history, 4) cognitive test scores, and 5) clinical observation and judgment (see Dawson et al., 2004 for further details). These participants were tested again at ages 6, 9, and 13-15 years. Forty-six participants from the original cohort were re-contacted and invited to participate in this current study. The remaining 26 were not contacted because they were already recruited for another study. Of the 46 participants contacted, 4 had moved out of state, 2 were not interested in participating, 1 could not be scheduled, 25 did not respond to phone calls or emails, and 2 did not meet our eligibility criteria of being able to speak in 3-word phrases. Twelve ASD participants from the original cohort were enrolled in the study and tested in their 20s (see Table 1 for sample demographics).

**Table 1.**
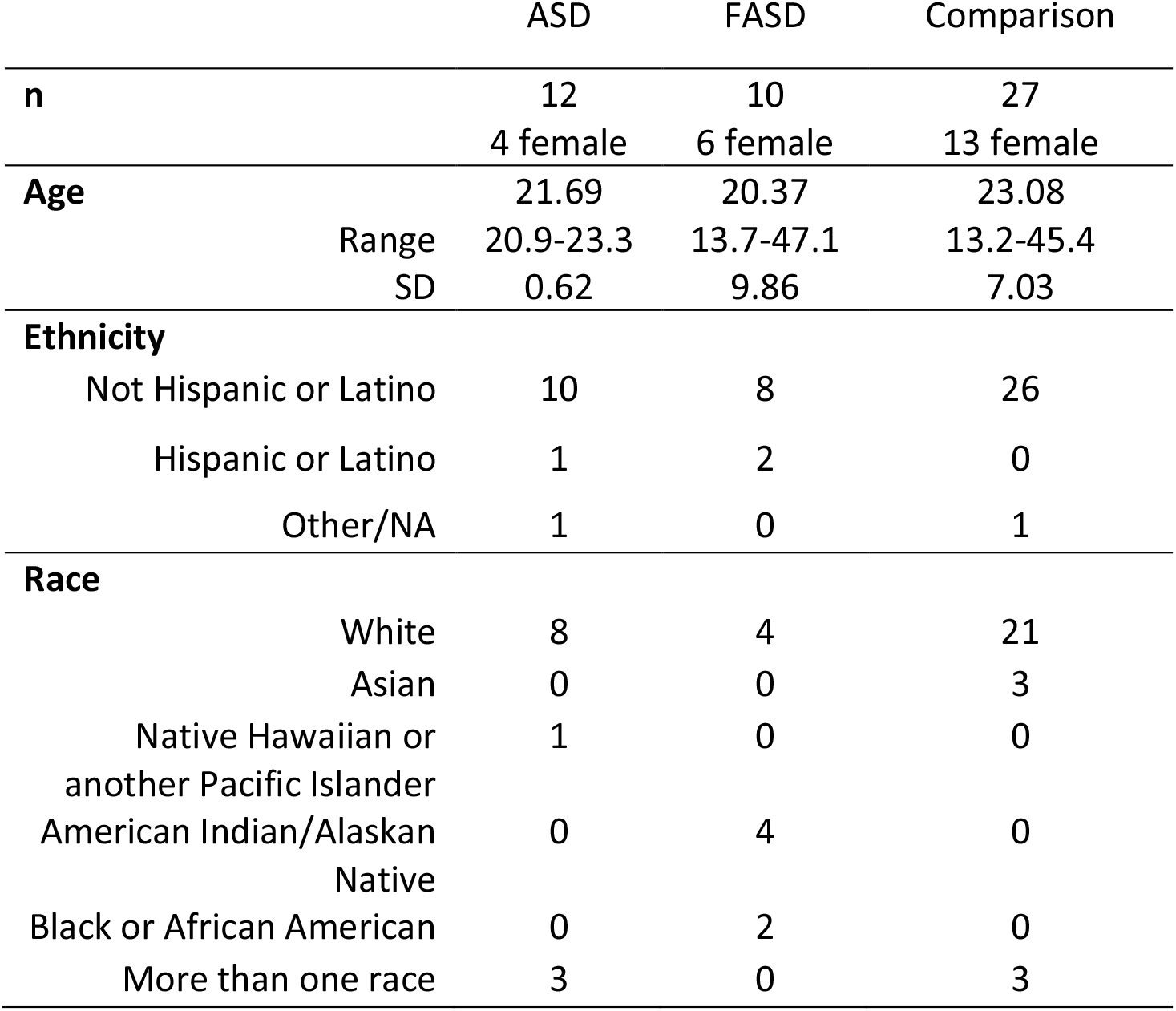
Sample Demographics

Ten participants with FASD were recruited from the University of Washington Fetal Alcohol Syndrome Diagnostic and Prevention Network (FAS DPN). All patients in the FAS DPN had a diagnosis of FASD as determined by the 4-Digit Diagnostic Code (Astley, 2004, 2013), which is an interdisciplinary approach to diagnosis guided by the magnitude of expression of the four features of FAS: growth deficiency, FAS facial phenotype, CNS structural/functional abnormalities, and prenatal alcohol exposure. Two participants had a diagnosis of fetal alcohol syndrome, two with a diagnosis of static encephalography/alcohol exposed, and six with a diagnosis of neurobehavioral disorder/alcohol exposed (see Table 2 for speech perception thresholds and cognitive ability by FASD diagnoses). Nine of ten FASD participants were adolescent/young adults who were teenagers and in their 20s; however, one adult in their 40s was also included in the study.

**Table 2:** Multitalker speech perception thresholds (TMR in dB) and WASI-II Full Scale IQ (FSIQ-4) by FASD diagnoses for participants in the FASD group (Astley, 2004, 2013).

|  | n | TMR | WASI-II FSIQ-4 |
| --- | --- | --- | --- |
| Fetal Alcohol Syndrome | 2 | 4.5(0.34) | 70.00(10.00) |
| Static Encephalography/Alcohol Exposed | 2 | 3.53(2.75) | 97.00(10.00) |
| Neurobehavioral Disorder/Alcohol Exposed | 6 | -0.16(1.08) | 106.00(6.37) |

*Note:* Mean ± SE shown in brackets.

The Comparison group consisted of 27 neurotypical participants. For each participant in the ASD and FASD groups, one age- and sex-matched participant was recruited as a pair matched comparison subject. Age matching in comparison participants was ± 1 year except the participant in their 40s in the FASD group was matched within 2 years. A one-way analysis of variance confirmed that there was no significant difference in age between groups (F(2,46) = 0.611, *p* = 0.547). Participants were also matched on sex due to the higher incidence of males with a diagnosis of ASD. Besides the pair matched comparison subjects, there were also an additional five neurotypical participants tested. Comparison group participants all reported no history of cognitive, developmental, or other health concerns. Two participants in the Comparison group had a diagnosis of ADHD but were included because there were individuals with ADHD in both ASD and FASD groups.

### Stimuli

Sentence stimuli were from the Coordinate Response Measure (CRM) corpus (Bolia et al., 2000), which consists of 256 sentences of the form “Ready (CALLSIGN) go to (COLOR) (NUMBER) now.” There are eight possible call signs (Arrow, Baron, Charlie, Eagle, Hopper, Laker, Ringo, Tiger), four colors (red, green, white, blue) and the numbers 1-8. Two male and two female speakers speaking all combinations of the call signs, colors, and numbers were included in the stimuli. Head-related transfer functions (HRTFs) from the CIPIC HRTF database (Algazi et al., 2001), were resampled to 24,414 to match the stimulus sampling rate then convolved with the target and masker sentences to simulate the source locations tested in this experiment: the target stream to be attended at 0° azimuth and the two spatially separated masking speech streams at ±45° azimuth. The target talker was always male, while the two maskers were either male/male or female/female. Sentences were randomly selected on each presentation.

### Procedures

The following measures were obtained over the course of several visits and as part of a larger study that included additional neurophysiological and behavioral measures. Written informed consent was obtained from all participants or their Legally Authorized Representative for participants under 16 years of age. All methods were performed in accordance with the relevant guidelines and regulations of protocols reviewed and approved by the Institutional Review Board at the University of Washington where the research was conducted. Participants were provided with monetary compensation for their time.

### ADOS-2

The ADOS-2 (Lord et al., 2012), a measure of autism symptom severity, was administered to all participants at the time of testing to confirm group inclusion. All participants in the ASD group received a classification of autism or autism spectrum based on their performance on the ADOS-2 while all participants in the FASD or Comparison received a classification of non-spectrum.

### WASI-II

While there are many cognitive processes that potentially contribute to multitalker speech perception, we administered a test of cognitive ability as a proxy measure. All four subtests of the Weschler Abbreviated Scale of Intelligence – Second Edition (WASI-II; Weschler, 2011): Block Design, Vocabulary, Matrix Reasoning, and Similarities was administered to each participant and considered as an overall estimate of their general cognitive functioning. The Verbal Composite Index (VCI) score was computed from the Vocabulary and Similarities subtests while the Perceptual Reasoning Index (PRI) scores were computed from the Block Design and Matrix Reasoning subtests.

### Audiological Screening

Typical hearing was an inclusion criterion to participate in this study. To ensure clinically normal hearing thresholds, all participants were required to pass an audiometric screen (≤ 20dB hearing level at octave frequencies between 250 and 8000 Hz) as well as a distortion product otoacoustic emission (DPOAE) screen. For the DPOAE screening, DPOAE growth functions were estimated at the distortion frequency of 2f1-f2. The frequency of the f2 primary tone was 4 kHz and the frequency and level of the f1 tone were varied according to formula in Johnson et al. (2006). One participant with ASD was excluded from this study because they failed the audiometric screening. One Comparison group participant was excluded from this study due to contraindication for magnetoencephalography recording.

### Multitalker Adaptive Threshold Task

To assess multi-talker speech perception, participants listened to three simultaneous sentences and were asked to identify the color and number spoken by the target talker, who was always situated in front. The two competing talkers, who we refer to as “maskers,” were presented off to the side (Fig. 1). At the start of the task, the target talker is the loudest with the competing maskers presented at a much lower sound level. To increase task difficulty, we increased the sound level of the competing maskers adaptively to obtain a speech perception threshold for each participant.

**Fig. 1.**
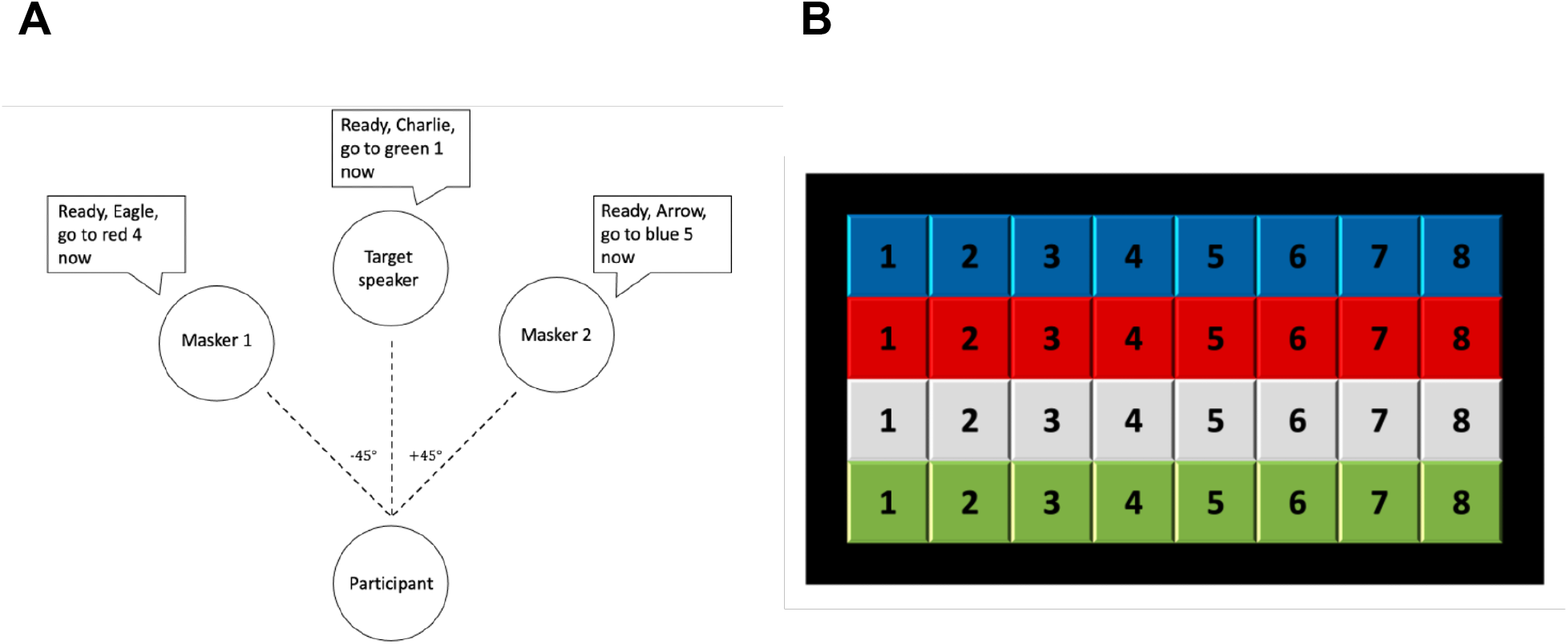
Schematic diagram of trial configuration and response panel. **A)** Participants heard three simultaneous sentences, with the target talker in front at 0° azimuth presented with two spatially separated simultaneous competing talkers, “maskers”, on each side at ±45° azimuth. **B)** Participants were asked to identify via mouse click the color and number spoken by the target talker on a computer screen.

These experimental procedures closely follow those outlined in Gallun et al. (2013), except that stimuli were presented via insert ear tips in this study as opposed to in sound field. Participants listened to three simultaneous sentences from the Coordinate Response Measure corpus (Bolia et al., 2000), which consists of 256 sentences of the form “Ready (CALLSIGN) go to (COLOR) (NUMBER) now.” The target stream to be attended was always positioned in front at 0° azimuth, with two spatially separated masking speech streams at ±45° azimuth. The goal was to attend to one of the three CRM sentences, identified by the callsign “Charlie,” and correctly identify the color and number spoken by the target talker. Speech perception thresholds, in terms of target-to-masker ratios (TMRs), were estimated using a one-up-one-down procedure, to estimate 50% correct (Levitt, 1971); the target level was fixed at 40 dB SPL and the masker levels were adaptively varied. The starting level for the masker sentences was 40 dB and they were changed in level by 5 dB for the first three reversals and then 1 dB for six additional reversals. Thresholds were estimated to be the geometric mean of the last six reversals. TMR is expressed as the level difference (dB) between the target and the two competing talkers, where a positive TMR indicated that the target had to be louder than the competing talkers, and a negative TMR indicated that the target talker would be quieter than the competing talkers. Thus, TMR scores in the positive range reflect poor speech perception performance, meaning that the listener needed the target talker to be louder than the maskers to accurately perceive the target talker’s speech. Each participant completed five adaptive tracks; the worst run (i.e., highest threshold value) was dropped and the best four runs were averaged.

To ensure audibility of the target sentence and to demonstrate the ability to perform the task, participants had to obtain 80% correct (8 out of 10 trials) on a training run where the target sentences were presented in quiet before testing began. If participants were not able to reach obtain 80% correct on this speech perception task in quiet after a maximum of three attempts, they were excluded from the study; no participants were excluded for failing training. Feedback was provided for correct and incorrect trials only during the training run(s) and not during the actual adaptive tracks.

All auditory stimuli were presented via Etymotic ER-2 insert earphones in a sound-attenuated booth. Participant responses were obtained using a computer located on a table in front of the listener. The listener initiated each trial and indicated the color and number keywords associated with the target callsign “Charlie” by clicking a virtual button on a grid of color and number targets. All participants completed the five runs in a single session of about 30 minutes. All procedures were conducted according to protocols approved by the University of Washington Institutional Review Board.

### Statistical Analysis

To test the main hypothesis that cognitive deficits are correlated with poor speech perception thresholds among young adults with typical hearing, a simple linear regression was used to investigate whether cognitive ability (WASI-II FSIQ-4) was significantly correlated with speech perception thresholds (TMR). Two further secondary analyses were conducted to assess the stability of this relationship. A simple linear regression was used to investigate whether cognitive ability (WASI-II FSIQ-4) was significantly correlated with speech perception thresholds (TMR) within each group. Next, to investigate the relationship between verbal and non-verbal abilities, paired sample t-tests were first used to test for differences in average VCI and PRI scores within each diagnostic group. A simple linear regression was then used to investigate the relationship between verbal (VCI) or non-verbal ability (PCI) and speech perception (TMR). All tests were evaluated against at two-tailed *p*<0.05 level of significance. Pair matching was conducted during the recruitment of the Comparison group participants to achieve a balance in age and sex between the study groups. This pair matching was not maintained in the statistical analyses because preliminary analyses conducted using pair matched linear regression with ASD and FASD groups and their respective comparison groups revealed no effect of age; thus, all 27 Comparison participants were combined into one group. The total sample size of 49 was sufficient to detect the relationship between speech perception and cognitive ability using a one-sided test with a significance level of 0.025 to test whether the observed slope of -0.68 is greater than zero with a power (1-β) of >0.99.

## Results

### Multitalker speech perception as a function of IQ

WASI-II Full Scale Intelligent Quotient-4 (FSIQ-4) scores were correlated with speech perception thresholds in the overall sample (Fig. 2A; linear regression, *β* = −0.68, *t* = −6.29, *p* < 0.0001, *R*^2^ = 0.46, η^2^ = 0.46); As cognitive ability decreased, speech perception performance (the ability of the listener to selectively attend to one talker in the presence of maskers) also decreased. We conducted two further analyses to assess the stability of this relationship. First, we investigated whether the relationship between WASI-II FSIQ-4 scores and speech perception thresholds held within each diagnostic group. Indeed, WASI-II FSIQ-4 score was found to be significantly correlated with speech perception thresholds in all three groups (Fig. 2B-D; linear regression, ASD, n=12 : *β* = −0.77, *t* = −3.85, *p* = 0.003, *R*^2^ = 0.60, *R*^2^ = 0.60 ; Comparison, n=27: *β* = −0.39, *t* = −2.10, *p* = 0.046, *R*^2^ = 0.15, η^2^ = 0.15 ; FASD, n=10: *β* = −0.67, *t* = −2.57, *p* = 0.033, *R*^2^ = 0.45, η^2^ = 0.45).

**Fig. 2.**
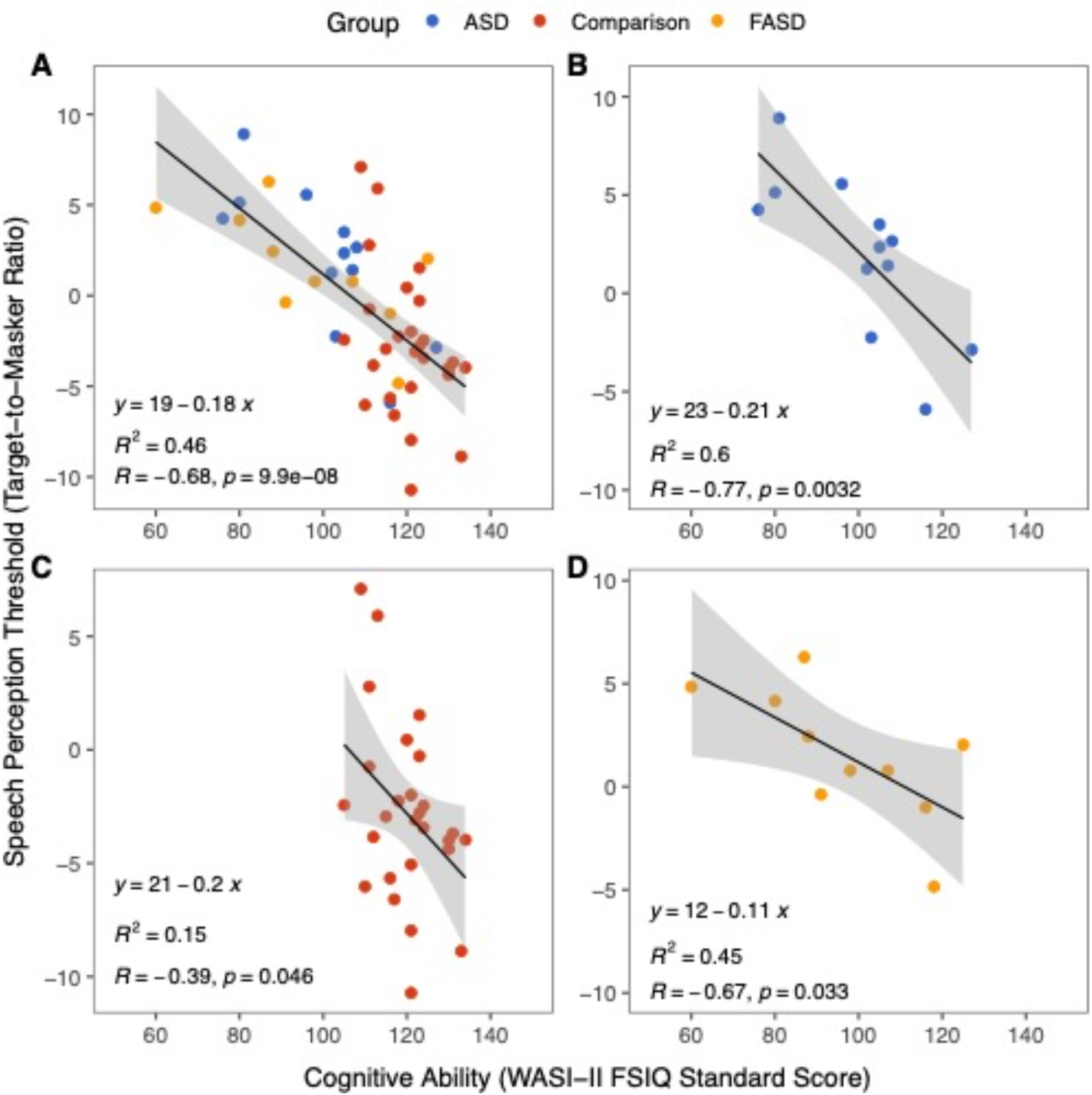
Speech perception performance improves with higher cognitive ability. **A)** Individual speech perception thresholds (TMR at 50% correct) as a function of cognitive ability (WASI-II FSIQ-4), with ASD group in blue, Comparison group in red, and FASD group in orange. Negative TMRs reflect better speech perception performance. WASI-II FSIQ-4 was correlated with TMR in the overall sample. **B-D)** Speech perception thresholds as a function of cognitive ability for the ASD (B), Comparison (C), and FASD (D) groups. Individual data points shown with solid circles. WASI-II FSIQ-4 was correlated with TMR in ASD, Comparison, and FASD groups.

### Verbal and non-verbal abilities

Next, we investigated the relationship between verbal and non-verbal abilities by separating the WASI-FSIQ-4 into the Verbal Comprehension Index score (VCI) and the Perceptual Reasoning Index score (PCI). We started by conducting paired samples t-tests to compare average VCI and PRI scores within each diagnostic group. We did not observe a difference between the two scores in any of the three groups (ASD paired *t*(11) = −1.35, *p* = 0.21; Comparison paired t(26) = −1.67, *p* = 0.11; FASD paired t(9) = 0.89, *p* = 0.4). Furthermore, both WASI-II VCI (Fig. 3A, linear regression, *β* = −0.56, *t* = −4.66, *p* < .0001, *R*^2^ = 0.32, η^2^ = 0.32) and WASI-II PRI were correlated with speech perception performance in the overall sample (Fig. 3B, linear regression, *β* = −0.63, *t* = −5.51, *p* < 0.0001, *R*^2^ = 0.39, η^2^ = 0.39). The higher the VCI and PRI scores, the better the speech perception performance, showing that the relationship holds for both verbal and nonverbal abilities.

**Fig. 3.**
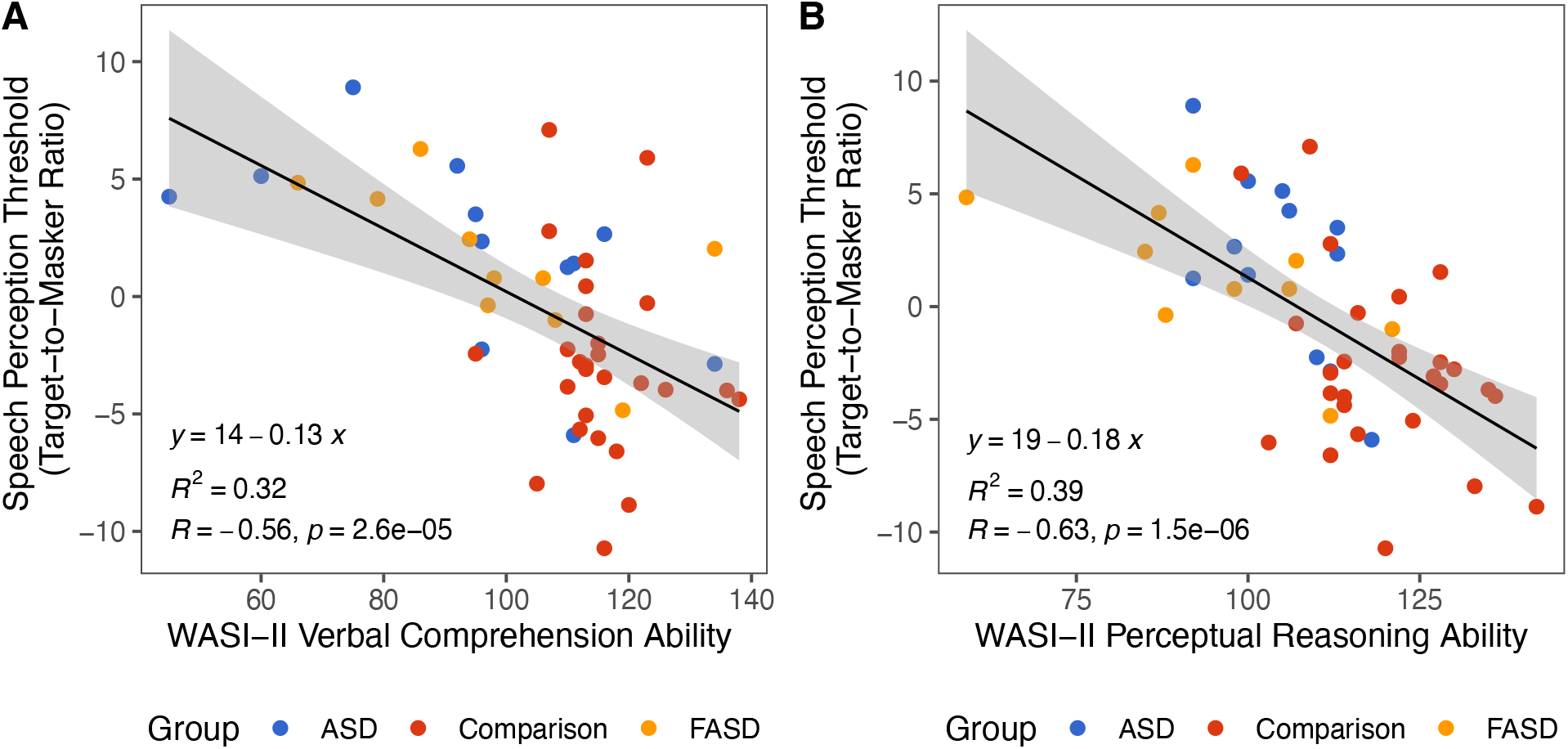
Speech perception performance correlated with both verbal and nonverbal abilities. **A)** Verbal comprehension ability (WASI-II VCI score) as a function of multitalker speech perception performance (TMR). Individual data points shown with solid circles. WASI-II VCI was significantly correlated with TMR. **B)** Perceptual reasoning ability (WASI-II PRI score) as a function of multitalker speech perception performance (TMR). Individual data points shown with solid circles. WASI-II PRI was significantly correlated with TMR.

## Discussion

We observed an association between cognitive ability and multitalker speech perception thresholds; as cognitive ability increased, multitalker speech perception thresholds improved. This relationship persisted even when we accounted for verbal and non-verbal abilities as two of the subtests on the WASI-II, Vocabulary and Similarities, required verbal responses while two of the subtests, Block Design and Matrix Reasoning did not.

On average, participants in the ASD and FASD groups required the target speaker to be louder than the two competing talkers, as reflected by positive target-to-masker ratios, in order to adequately attend to the target talker. In contrast, individuals in the Comparison group were able to selectively attend to the target talker even when competing voices were louder. These results are consistent with prior studies showing worse speech perception in autistic adults and children (Alcántara et al., 2004; DePape et al., 2012; Schelinski & von Kriegstein, 2020). They are also consistent with self/caregiver reports of auditory processing abnormalities including difficulty listening to speech under noisy conditions in individuals with FASD and ASD (Ben-Sasson et al., 2009; Church & Gerkin, 1988; Franklin et al., 2008; Gabriels et al., 2008; Green et al., 2016; McLaughlin et al., 2019). However, one limitation of the study is that we did not have the statistical power to test between group differences. Future studies with larger sample sizes will be required to further investigate multitalker speech perception differences between FASD, ASD, and comparison groups, while controlling for IQ.

That the relationship between cognitive abilities and multitalker speech perception was observed in all participant groups, including the neurotypical Comparison group, provides evidence that cognitive processes play an integral role in listening under complex conditions. If multitalker speech perception were driven primarily by auditory processes, we would not expect to see an effect of cognitive ability on speech perception thresholds in neurotypical adults with typical hearing. Moreover, degradation in the auditory peripheral encoding of the input speech mixture, such as in individuals with hearing loss, is associated with deficits in all subsequent processing stages including the segregation and grouping of sounds as well as complex processing of speech specific information such as semantics and syntax (Dai et al., 2018; Grimault et al., 2001; Shinn-Cunningham & Best, 2008). Instead, our results show that impairments in multitalker speech perception do not have to originate in the auditory periphery, as all participants in this study had clinically normal hearing thresholds. As impairments in multitalker speech perception do not have to originate in the auditory periphery, there are important implications for both conceptual models of multitalker speech perception as well as clinical implications for neurodivergent individuals. Whereas listening in noisy environments is known to be difficult for individuals who are hard of hearing, with hearing assistive technology and auditory rehabilitation therapy often employed to address this challenge, our data suggest that individuals with lower cognitive ability may also have trouble listening under complex acoustic conditions. This suggests that those children who are cognitively challenged and the furthest behind their peers will have the most need to learn in noisy classrooms but, may also have the most trouble listening. Consequently, future studies are warranted to further investigate the need for targeted assessment and intervention for speech perception difficulties among neurodivergent individuals living in noisy real-world conditions.

It is unclear from this current study which stages of multitalker speech perception are affected by impairments in cognition. In a related study investigating stream segregation and selective attention, we did not observe an effect of cognitive ability in the ASD or Comparison group (Emmons et al., 2021). One possible explanation for these divergent findings is the difference in cognitive load between the tasks (Peelle, 2018). In Emmons et al. 2021 (Emmons et al., 2021) we presented a dual stream selective attention task with two talkers who each spoke two words. In this current paradigm, there are three simultaneous talkers speaking full sentences, that requires greater listening effort. For individuals with higher ability, this increased cognitive load may not have a discernable effect on complex listening. For individuals with lower ability, however, the effort associated with segregating and selectively listening in a multitalker situation may be sufficient to reduce their ability to remember a color and a number that was spoken by the target speaker. This may be especially true for individuals with FASD, who often experience deficits in working memory. Moreover, this conjecture is consistent with the fact that all participants in this study, regardless of ability, were able to perform the multitalker speech perception task with positive TMRs when the target talker was louder than the competing talkers.

There are several important limitations to consider when interpretating findings from the present study. First, Comparison group participants in our sample have a limited IQ range. While a significant relationship between cognitive ability and multitalker speech perception is observed in this group for average to above average IQs, future studies should aim to replicate these findings in neurotypical adults with lower IQs. Second, future studies should evaluate the possibility that attention deficits could impact multitalker speech perception. There is evidence that the selective enhancement of relevant speech is influenced by an attention-mediated feedback loop (Ahveninen et al., 2011; Bronkhorst, 2015; Ding & Simon, 2012; O’Sullivan et al., 2015; Woldorff & Hillyard, 1991). Although we did not assess attention in this study, we obtained a medical history from the participant or their parent/guardian prior to participation and found no evidence for a relationship between report of ADHD and performance on out multitalker speech perception task. However, direct assessment of attentional abilities is needed to further evaluate this potential contributor. Finally, future studies with larger cohorts of ASD and FASD participants are needed to further investigate individual variability in these heterogeneous populations. However, our results provide novel evidence that listening in complex situations may be impaired in individuals with lower IQ.

In conclusion, in this investigation of multitalker speech perception in young, typical hearing, neurodivergent individuals, we found a highly significant relationship between cognitive ability and multitalker speech perception, that held for each diagnostic group individually. This observation has both theoretical and clinical importance, showing that deficits in complex listening do not have to originate in the peripheral encoding of sound as it is commonly assumed; instead, cognitive abilities could also have significant influences. From a clinical perspective, these findings warrant further investigation into listening under complex, real-world environments to support learning and communication for neurodivergent individuals with lower cognitive ability.

## Data Availability

All data produced in the present study are available upon reasonable request to the authors

## Acknowledgements

We thank Erick Gallun for helpful discussions relating to this project.

## Data Availability

All data and analysis code are available from corresponding author upon request.

## References

Ahveninen, J., Hämäläinen, M., Jääskeläinen, I. P., Ahlfors, S. P., Huang, S., Lin, F.-H., Raij, T., Sams, M., Vasios, C. E., & Belliveau, J. W. (2011). Attention-driven auditory cortex short-term plasticity helps segregate relevant sounds from noise. Proceedings of the National Academy of Sciences of the United States of America, 108(10), 4182–4187. https://doi.org/10.1073/pnas.1016134108

Alcántara, J. I., Weisblatt, E. J. L., Moore, B. C. J., & Bolton, P. F. (2004). Speech-in-noise perception in high-functioning individuals with autism or Asperger’s syndrome. Journal of Child Psychology and Psychiatry, 45(6), 1107–1114. https://doi.org/10.1111/j.1469-7610.2004.t01-1-00303.x

Algazi, V. R., Duda, R. O., Thompson, D. M., & Avendano, C. (2001). The CIPIC HRTF database. Proceedings of the 2001 IEEE Workshop on the Applications of Signal Processing to Audio and Acoustics (Cat. No.01TH8575), 99–102. https://doi.org/10.1109/ASPAA.2001.969552

American Psychiatric Association. (1994). Diagnostic and statistical manual of mental disorders: DSM-IV-TR (4th ed.). author.

American Psychiatric Association (Ed.). (2013). Diagnostic and statistical manual of mental disorders: DSM-5. American Psychiatric Association.

Astley, S. (2004). Diagnostic guide for fetal alcohol spectrum disorders: The 4-digit diagnostic code.

Astley, S. (2013). Validation of the fetal alcohol spectrum disorder (fasd) 4-digit diagnostic code. Journal of Population Therapeutics and Clinical Pharmacology, 20(3), Article 3. https://jptcp.com

Ben-Sasson, A., Hen, L., Fluss, R., Cermak, S. A., Engel-Yeger, B., & Gal, E. (2009). A meta-analysis of sensory modulation symptoms in individuals with autism spectrum disorders. Journal of Autism and Developmental Disorders, 39(1), 1–11. https://doi.org/10.1007/s10803-008-0593-3

Best, V., Gallun, F. J., Ihlefeld, A., & Shinn-Cunningham, B. G. (2006). The influence of spatial separation on divided listening. The Journal of the Acoustical Society of America, 120(3), 1506–1516. https://doi.org/10.1121/1.2234849

Birch, J. (2003). Congratulations! It’s asperger syndrome. Jessica Kingsley Publishers.

Bizley, J. K., & Cohen, Y. E. (2013). The what, where and how of auditory-object perception. Nature Reviews. Neuroscience, 14(10), 693–707. https://doi.org/10.1038/nrn3565

Bolia, R., Nelson, W., Ericson, M., & Simpson, B. (2000). A speech corpus for multitalker communications research. The Journal of the Acoustical Society of America, 107, 1065– 1066. https://doi.org/10.1121/1.428288

Bregman, A. S. (1990). Auditory scene analysis: The perceptual organization of sound. MIT Press.

Bronkhorst. (2015). The cocktail-party problem revisited: Early processing and selection of multi-talker speech. Attention, Perception, & Psychophysics, 77(5), 1465–1487. https://doi.org/10.3758/s13414-015-0882-9

Bronkhorst, A. (2000). The cocktail party phenomenon: A review of research on speech intelligibility in multiple-talker conditions. Acta Acustica United with Acustica, 86(1), 117–128.

Brungart, D. S. (2001). Informational and energetic masking effects in the perception of two simultaneous talkers. The Journal of the Acoustical Society of America, 109(3), 1101– 1109. https://doi.org/10.1121/1.1345696

Church, M. W., & Gerkin, K. P. (1988). Hearing disorders in children with fetal alcohol syndrome: Findings from case reports. Pediatrics, 82(2), 147–154. https://doi.org/10.1542/peds.82.2.147

Dai, L., Best, V., & Shinn-Cunningham, B. G. (2018). Sensorineural hearing loss degrades behavioral and physiological measures of human spatial selective auditory attention. Proceedings of the National Academy of Sciences, 115(14), E3286–E3295. https://doi.org/10.1073/pnas.1721226115

Davis, M. H., & Johnsrude, I. S. (2007). Hearing speech sounds: Top-down influences on the interface between audition and speech perception. Hearing Research, 229(1), 132–147. https://doi.org/10.1016/j.heares.2007.01.014

Dawson, G., Toth, K., Abbott, R., Osterling, J., Munson, J., Estes, A., & Liaw, J. (2004). Early social attention impairments in autism: Social orienting, joint attention, and attention to distress. Developmental Psychology, 40(2), 271–283. https://doi.org/10.1037/0012-1649.40.2.271

DePape, A.-M. R., Hall, G. B. C., Tillmann, B., & Trainor, L. J. (2012). Auditory processing in high-functioning adolescents with autism spectrum disorder. PLOS ONE, 7(9), e44084. https://doi.org/10.1371/journal.pone.0044084

Ding, N., & Simon, J. Z. (2012). Emergence of neural encoding of auditory objects while listening to competing speakers. Proceedings of the National Academy of Sciences, 109(29), 11854–11859. https://doi.org/10.1073/pnas.1205381109

Emmons, K. A., KC Lee, A., Estes, A., Dager, S., Larson, E., McCloy, D. R., St John, T., & Lau, B. K. (2021). Auditory attention deployment in young adults with autism spectrum disorder. Journal of Autism and Developmental Disorders. https://doi.org/10.1007/s10803-021-05076-8

Franklin, L., Deitz, J., Jirikowic, T., & Astley, S. (2008). Children with fetal alcohol spectrum disorders: Problem behaviors and sensory processing. The American Journal of Occupational Therapy: Official Publication of the American Occupational Therapy Association, 62(3), 265–273. https://doi.org/10.5014/ajot.62.3.265

Gabriels, R. L., Agnew, J. A., Miller, L. J., Gralla, J., Pan, Z., Goldson, E., Ledbetter, J. C., Dinkins, J. P., & Hooks, E. (2008). Is there a relationship between restricted, repetitive, stereotyped behaviors and interests and abnormal sensory response in children with autism spectrum disorders? Research in Autism Spectrum Disorders, 2(4), 660–670. https://doi.org/10.1016/j.rasd.2008.02.002

Gallun, F. J., & Best, V. (2020). Age-related changes in segregation of sound sources. In K. S. Helfer, E. L. Bartlett, A. N. Popper, & R. R. Fay (Eds.), Aging and Hearing: Causes and Consequences (pp. 143–171). Springer International Publishing. https://doi.org/10.1007/978-3-030-49367-7_7

Gallun, F. J., Diedesch, A. C., Kampel, S. D., & Jakien, K. M. (2013). Independent impacts of age and hearing loss on spatial release in a complex auditory environment. Frontiers in Neuroscience, 7. https://doi.org/10.3389/fnins.2013.00252

Grandin, T. (2006). Thinking in pictures: And other reports from my life with autism. Vintage.

Green, D., Chandler, S., Charman, T., Simonoff, E., & Baird, G. (2016). Brief report: Dsm-5 sensory behaviours in children with and without an autism spectrum disorder. Journal of Autism and Developmental Disorders, 46(11), 3597–3606. https://doi.org/10.1007/s10803-016-2881-7

Grimault, N., Micheyl, C., Carlyon, R. P., Arthaud, P., & Collet, L. (2001). Perceptual auditory stream segregation of sequences of complex sounds in subjects with normal and impaired hearing. British Journal of Audiology, 35(3), 173–182. https://doi.org/10.1080/00305364.2001.11745235

Groen, W. B., van Orsouw, L., Huurne, N. ter, Swinkels, S., van der Gaag, R.-J., Buitelaar, J. K., & Zwiers, M. P. (2009). Intact spectral but abnormal temporal processing of auditory stimuli in autism. Journal of Autism and Developmental Disorders, 39(5), 742–750. https://doi.org/10.1007/s10803-008-0682-3

Haesen, B., Boets, B., & Wagemans, J. (2011). A review of behavioural and electrophysiological studies on auditory processing and speech perception in autism spectrum disorders. Research in Autism Spectrum Disorders, 5(2), 701–714. https://doi.org/10.1016/j.rasd.2010.11.006

Humes, L. E., Busey, T. A., Craig, J., & Kewley-Port, D. (2013). Are age-related changes in cognitive function driven by age-related changes in sensory processing? Attention, Perception & Psychophysics, 75(3), 508–524. https://doi.org/10.3758/s13414-012-0406-9

Johnson, T. A., Neely, S. T., Garner, C. A., & Gorga, M. P. (2006). Influence of primary-level and primary-frequency ratios on human distortion product otoacoustic emissions. The Journal of the Acoustical Society of America, 119(1), 418–428. https://doi.org/10.1121/1.2133714

Klin, A. (1993). Auditory brainstem responses in autism: Brainstem dysfunction or peripheral hearing loss? Journal of Autism and Developmental Disorders, 23(1), 15–35. https://doi.org/10.1007/BF01066416

Levitt, H. (1971). Transformed up-down methods in psychoacoustics. The Journal of the Acoustical Society of America, 49(2B), 467–477. https://doi.org/10.1121/1.1912375

Lord, C., Rutter, M., DiLavore, P. C., Risi, S., Gotham, K., & Bishop, S. L. (2012). Autism diagnostic observation schedule, 2nd. Western Psychological Services.

Lord, C., Rutter, M., Goode, S., Heemsbergen, J., Jordan, H., Mawhood, L., & Schopler, E. (1989). Autism diagnostic observation schedule: A standardized observation of communicative and social behavior. Journal of Autism and Developmental Disorders, 19(2), 185–212. https://doi.org/10.1007/BF02211841

Lord, C., Rutter, M., & Le Couteur, A. (1994). Autism Diagnostic Interview-Revised: A revised version of a diagnostic interview for caregivers of individuals with possible pervasive developmental disorders. Journal of Autism and Developmental Disorders, 24(5), 659– 685. https://doi.org/10.1007/BF02172145

McLaughlin, S. A., Thorne, J. C., Jirikowic, T., Waddington, T., Lee, A. K. C., & Astley, H. S. J. (2019). Listening difficulties in children with fetal alcohol spectrum disorders: More than a problem of audibility. Journal of Speech, Language, and Hearing Research, 62(5), 1532–1548. https://doi.org/10.1044/2018_JSLHR-H-18-0359

Michalka, S. W., Kong, L., Rosen, M. L., Shinn-Cunningham, B. G., & Somers, D. C. (2015). Short-term memory for space and time flexibly recruit complementary sensory-biased frontal lobe attention networks. Neuron, 87(4), 882–892. https://doi.org/10.1016/j.neuron.2015.07.028

Moore, B. C. J. (2003). Speech processing for the hearing-impaired: Successes, failures, and implications for speech mechanisms. Speech Communication, 41(1), 81–91. https://doi.org/10.1016/S0167-6393(02)00095-X

Obler, L. K., Fein, D., Nicholas, M., & Albert, M. L. (1991). Auditory comprehension and aging: Decline in syntactic processing. Applied Psycholinguistics, 12(4), 433–452. https://doi.org/10.1017/S0142716400005865

O’Connor, K. (2012). Auditory processing in autism spectrum disorder: A review. Neuroscience & Biobehavioral Reviews, 36(2), 836–854. https://doi.org/10.1016/j.neubiorev.2011.11.008

O’Sullivan, J. A., Power, A. J., Mesgarani, N., Rajaram, S., Foxe, J. J., Shinn-Cunningham, B. G., Slaney, M., Shamma, S. A., & Lalor, E. C. (2015). Attentional selection in a cocktail party environment can be decoded from single-trial eeg. Cerebral Cortex, 25(7), 1697– 1706. https://doi.org/10.1093/cercor/bht355

Peelle, J. E. (2018). Listening effort: How the cognitive consequences of acoustic challenge are reflected in brain and behavior. Ear and Hearing, 39(2), 204–214. https://doi.org/10.1097/AUD.0000000000000494

Peelle, J. E., Troiani, V., Grossman, M., & Wingfield, A. (2011). Hearing loss in older adults affects neural systems supporting speech comprehension. Journal of Neuroscience, 31(35), 12638–12643. https://doi.org/10.1523/JNEUROSCI.2559-11.2011

Rosenhall, U., Nordin, V., Sandström, M., Ahlsén, G., & Gillberg, C. (1999). Autism and hearing loss. Journal of Autism and Developmental Disorders, 29(5), 349–357. https://doi.org/10.1023/A:1023022709710

Schelinski, S., & von Kriegstein, K. (2020). Brief report: Speech-in-noise recognition and the relation to vocal pitch perception in adults with autism spectrum disorder and typical development. Journal of Autism and Developmental Disorders, 50(1), 356–363. https://doi.org/10.1007/s10803-019-04244-1

Shinn-Cunningham, B., & Best, V. (2008). Selective attention in normal and impaired hearing. Trends in Amplification, 12(4), 283–299. https://doi.org/10.1177/1084713808325306

Shinn-Cunningham, B., Best, V., & Lee, A. K. C. (2017). Auditory object formation and selection. In J. C. Middlebrooks, J. Z. Simon, A. N. Popper, & R. R. Fay (Eds.), The Auditory System at the Cocktail Party (pp. 7–40). Springer International Publishing. https://doi.org/10.1007/978-3-319-51662-2_2

Szymanski, C. A., Brice, P. J., Lam, K. H., & Hotto, S. A. (2012). Deaf children with autism spectrum disorders. Journal of Autism and Developmental Disorders, 42(10), 2027–2037. https://doi.org/10.1007/s10803-012-1452-9

Taljaard, D. s., Olaithe, M., Brennan-Jones, C. g., Eikelboom, R. h., & Bucks, R. s. (2016). The relationship between hearing impairment and cognitive function: A meta-analysis in adults. Clinical Otolaryngology, 41(6), 718–729. https://doi.org/10.1111/coa.12607

Tomchek, S. D., & Dunn, W. (2007). Sensory processing in children with and without autism: A comparative study using the short sensory profile. American Journal of Occupational Therapy, 61(2), 190–200. https://doi.org/10.5014/ajot.61.2.190

Van Gerven, P. W. M., & Guerreiro, M. J. S. (2016). Selective attention and sensory modality in aging: Curses and blessings. Frontiers in Human Neuroscience, 10, 147. https://doi.org/10.3389/fnhum.2016.00147

Weschler, D. (2011). Weschler abbreviated scale of intelligence, second edition (WASI-II). NCS Pearson.

Woldorff, M. G., & Hillyard, S. A. (1991). Modulation of early auditory processing during selective listening to rapidly presented tones. Electroencephalography and Clinical Neurophysiology, 79(3), 170–191. https://doi.org/10.1016/0013-4694(91)90136-R

